# Biopsy-Validated Deep Learning Mapping of Glioma Infiltration Beyond Contrast Enhancement Using Multiparametric MRI

**DOI:** 10.64898/2026.10.26.26366551

**Authors:** Robert T Wujek, Melissa A Prah, Kathleen M Schmainda

## Abstract

**Background:** Glioblastoma (GBM) exhibits diffuse tumor infiltration beyond contrast-enhancing margins into FLAIR-hyperintense brain tissue, where infiltrative tumor is not visible on conventional MRI. We developed and validated the infiltrative Tumor Burden (iTB) map, a deep learning biomarker that uses multiparametric MRI (mpMRI) to identify infiltrative tumor within these non-enhancing regions.

**Methods:** Preoperative mpMRI datasets from 80 patients with WHO grade II–IV gliomas and 24 patients with brain metastases were retrospectively analyzed. Imaging included pre- and post-contrast T1-weighted, T2-weighted, FLAIR, dynamic susceptibility contrast perfusion and diffusion MRI-derived maps (rCBV, rCBF, ADC). Spatially localized stereotactic biopsies from non-enhancing glioma regions provided histopathologic ground truth while peritumoral brain metastasis regions served as biologically grounded non-tumor controls. A 3D convolutional neural network was trained on voxel-centered mpMRI patches using patient-level cross-validation and an independent test set, with data augmentation optimized using a greedy local search strategy.

**Results:** Model development included 121 biopsy-localized samples from 56 glioma patients and 104 non-tumor samples from 26 brain metastasis patients. Augmentation optimization significantly improved independent test-set performance, increasing accuracy from 74.5% to 85.5% and AUC from 0.859 to 0.942 (p = 0.008). Ensemble-derived voxel-wise iTB maps demonstrated low infiltration probabilities in brain metastases and extensive regions of elevated infiltration probability in glioblastoma, consistent with expected biology.

**Conclusions:** The iTB biomarker generates voxel-wise maps of tumor infiltration within non-enhancing FLAIR abnormalities. Validated against image-localized histopathology and independent testing, it achieved high diagnostic performance and may improve surgical planning, radiotherapy targeting, and assessment of glioma burden.

## 1.0 INTRODUCTION

Glioblastoma (GBM) is the most common and aggressive malignant brain tumor in adults, with a median overall survival of only 12-14 months despite intensive treatment, including maximal safe resection followed by concurrent radiation therapy and chemotherapy (1,2). A key factor underlying the poor prognosis of GBM is its highly invasive nature, combined with the inability to reliably identify tumor cells infiltrating the surrounding peritumoral brain tissue. The persistence of these infiltrating cells almost invariably leads to recurrence (3–7).

The clinical importance of this infiltration is highlighted by studies showing that survival may be improved through supramaximal resection – removal of not only the contrast-enhancing tumor visible on post-contrast T1-weighted MRI (T1+C), but also part of the surrounding non-enhancing tissue that appears hyperintense on FLAIR (Fluid Attenuated Inversion Recovery) images (8–10). However, a key challenge remains; conventional imaging cannot reliably distinguish between infiltrating tumor cells and vasogenic edema within these non-enhancing regions. This limitation underscores the need for an imaging biomarker that can accurately differentiate tumor infiltration from edema. Such a biomarker would enable more precise surgical planning and targeted radiation therapy and could also enhance treatment monitoring by providing a more accurate assessment of the extent of active tumor.

Several studies have employed multiparametric MRI (mpMRI) in conjunction with supervised machine learning to address this challenge (11–27), albeit with limited success in terms of performance and clinical translation (28,29). These studies commonly utilize a combination of conventional MRI sequences (T1, T1+C, T2, FLAIR) alongside advanced modalities such as diffusion-weighted imaging (DWI) and dynamic susceptibility contrast (DSC) perfusion MRI as predictive inputs with gross histopathology or radiographic assessment as ground truth outputs. Notably, substantial heterogeneity exists across studies in terms of dataset composition, tissue labeling strategies, and machine learning methods.

Although promising results have been reported, clinical translation remains limited by several key methodological challenges. A major issue is the difficulty of comparing models across studies, as performance metrics are reported inconsistently. Common measures such as accuracy, sensitivity, specificity, and AUC are not uniformly provided, and clinically meaningful metrics, including positive predictive value (PPV) and negative predictive value (NPV) are often omitted.

Many studies are also based on small sample sizes, increasing the risk of overfitting and limiting the robustness and generalizability of findings. In addition, model performance is often evaluated using cross-validation (CV) schemes without independent test cohorts (12,14,19,21,24,27), raising concerns about overestimation of real-world performance.

Ground-truth labeling presents another significant limitation. A substantial proportion of studies rely on indirect rather than image-localized histopathologic confirmation. Tumor presence is often inferred from surrogate markers such as increased cell density (17,19), which may also reflect non-tumor abnormalities; proximity to contrast-enhancing lesion boundaries (13); spatial and temporal correspondence with regions that later demonstrate tumor progression on imaging (11,12,14,16,23–26), or findings from delayed postmortem sampling (15).

Collectively, these limitations highlight the need for larger, well-validated studies incorporating independent test datasets and more precise ground truth definitions to improve the reliability and clinical applicability of predictive models.

In this study, we developed a novel 3D convolutional neural network (CNN) designed to leverage mpMRI for the identification of infiltrating tumor within non-enhancing brain tissue. Using a relatively large dataset with image-localized histopathology as the ground truth, the resulting infiltrative tumor burden (iTB) model generates voxel-wise probability maps of tumor infiltration within segmented non-enhancing regions.

A key strength of this study is the use of a comparatively large, histologically validated dataset comprising stereotactically acquired biopsy samples that were spatially matched to mpMRI obtained within a short time interval. Combined with virtual tissue samples derived from pathologically confirmed brain metastases, which are characteristically expansive and non-infiltrative, these data provided a rigorous reference standard for tissue labeling and model development. The size of the dataset further enabled the development and independent validation of the proposed iTB model through separate training and validation cohorts. To support robust model training, we also implemented a greedy local-search strategy to optimize data augmentation techniques specifically designed for this mpMRI framework, effectively expanding the training dataset and enhancing model performance.

## 2.0 METHODS

### 2.1 Patient Cohorts

The institutional review board at The Medical College of Wisconsin (MCW) approved this study, which accessed patient datasets from the MCW Brain and Spinal Cord Tissue Bank.

For the first patient cohort, patients with WHO grades II–IV gliomas were retrospectively identified for inclusion if they had undergone stereotactically-guided biopsy using the Stealth Station navigation system (Medtronic, Minneapolis, MN, USA), had intraoperative screen captures available for biopsy localization, and had undergone a comprehensive preoperative mpMRI examination, including T1-weighted (T1w), post-contrast T1-weighted (T1+C), T2-weighted (T2w), fluid-attenuated inversion recovery (FLAIR), dynamic susceptibility contrast (DSC) perfusion imaging, and diffusion-weighted imaging (DWI).

For the second patient cohort, patients with brain metastases were retrospectively identified for inclusion if they had undergone the same comprehensive preoperative mpMRI examination used in the glioma cohort and had a surgically confirmed diagnosis with documented primary tumor histology. This cohort was included to address sampling bias in the glioma dataset of stereotactic biopsies. Because these biopsies preferentially target imaging-suspicious regions, they tend to overrepresent tumor-involved tissue relative to non-tumor tissue.

Given the well-established imaging and pathological characteristics of brain metastases, namely, their predominantly expansile growth pattern with minimal infiltrative behavior (30), this cohort served as a non-infiltrative reference population. Its inclusion enabled a more balanced representation of tumor and non-tumor tissue within non-enhancing regions (13,14,31,32).

**Figure 1** summarizes inclusion and exclusion criteria for identifying glioma and brain metastases patients.

**Figure 1.**
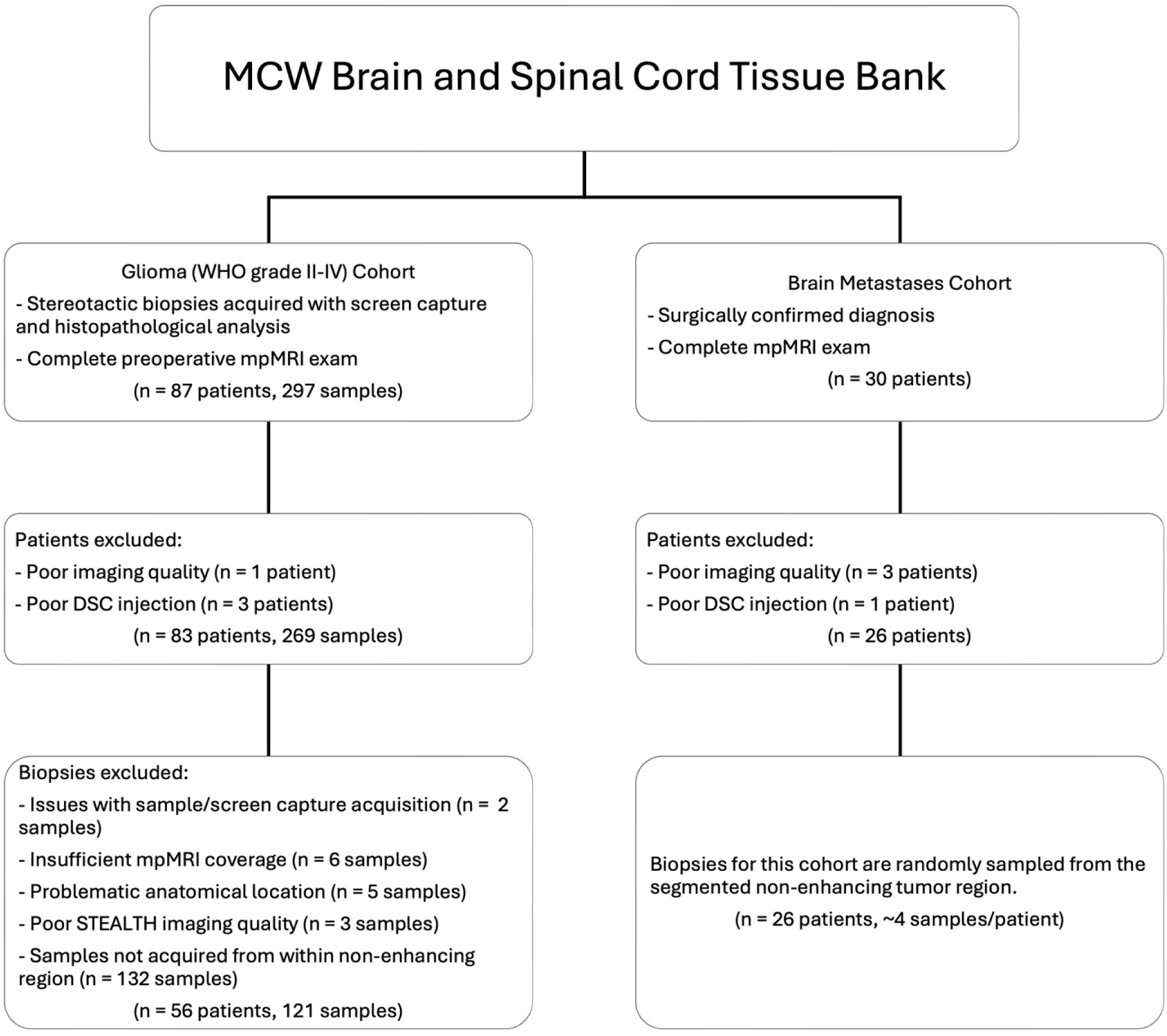
Study cohort selection and biopsy sample inclusion criteria. Patients from the MCW Brain and Spinal Cord Tissue Bank were assigned to the glioma (WHO grades II–IV) or brain metastases cohort based on diagnosis and imaging availability. Glioma biopsy samples were excluded for acquisition issues, insufficient mpMRI coverage, problematic anatomical location, poor STEALTH imaging quality, or sampling outside the non-enhancing lesion (NEL), yielding 121 eligible biopsy samples from 56 patients. The brain metastases cohort comprised 26 patients after exclusion for poor imaging quality or inadequate DSC injection. For this cohort, approximately four biopsy-sized regions of interest were randomly sampled per patient from the segmented NEL.

### 2.2 Multiparametric MRI Preprocessing

Multiparametric MRI (mpMRI) examinations were performed between 2009 and 2024 using 1.5T or 3T MRI systems from multiple vendors (GE Healthcare, Milwaukee, WI, USA; Siemens Healthineers, Munich, Germany) across multiple sites within the Froedtert-MCW healthcare system. Conventional imaging parameters were determined by routine clinical protocols and therefore varied across scanner platforms, acquisition sites, and over time. While this variability introduces heterogeneity in the data, it reflects real-world clinical conditions and may enhance the robustness and generalizability of machine learning models trained on these data.

The initial preprocessing step consisted of generating leakage-corrected (33,34) and standardized (35) relative cerebral blood volume (rCBV) and relative cerebral blood flow (rCBF) maps from the DSC acquisition using IB Neuro™ (Imaging Biometrics LLC, Elm Grove, WI), as well as apparent diffusion coefficient (ADC) maps from the DWI acquisition using b-values of 0 and 1000 s/mm² with IB Diffusion™ (Imaging Biometrics LLC, Elm Grove WI). N4ITK bias field correction (36) was applied to the anatomical sequences (T1w, T1+C, T2w, and FLAIR). All imaging volumes were then registered to the T1w volume and resampled to 1mm isotropic resolution using ANTsPy (37). A whole-brain mask was generated from the T1w volume using HD-BET (38) and applied to all registered volumes. All imaging volumes except standardized rCBV were intensity normalized using contralateral normal-appearing white matter (NAWM) as an internal reference region. The NAWM ROI was manually defined within the hemisphere contralateral to the lesion, and mean signal intensity within this region was used for per-channel normalization. This step provided a consistent patient-specific intensity reference for downstream processing.

For multi-examination cases, in which mpMRI and high-resolution presurgical imaging were acquired at separate time points but within close temporal proximity (<15 days), each examination was processed independently and later fused. Inter-examination registration was performed by aligning T1w images between moving and fixed examinations and applying a composite transformation (scan → T1_moving → T1_fixed) to all modalities in the moving examination.

Tumor segmentations were generated from the anatomical sequences using the BraTS (Brain Tumor Segmentation) Orchestrator (39). Automated lesion-specific segmentation models were used to generate initial segmentations, which were subsequently reviewed and manually adjusted in ITK-SNAP (40).

The final mpMRI dataset comprised T1w, T1+C, T2w, FLAIR, rCBV, rCBF, and ADC volumes, along with BraTS-derived tumor segmentation maps, all in a common T1w space. The full preprocessing pipeline is illustrated in **Figure 2**.

**Figure 2.**
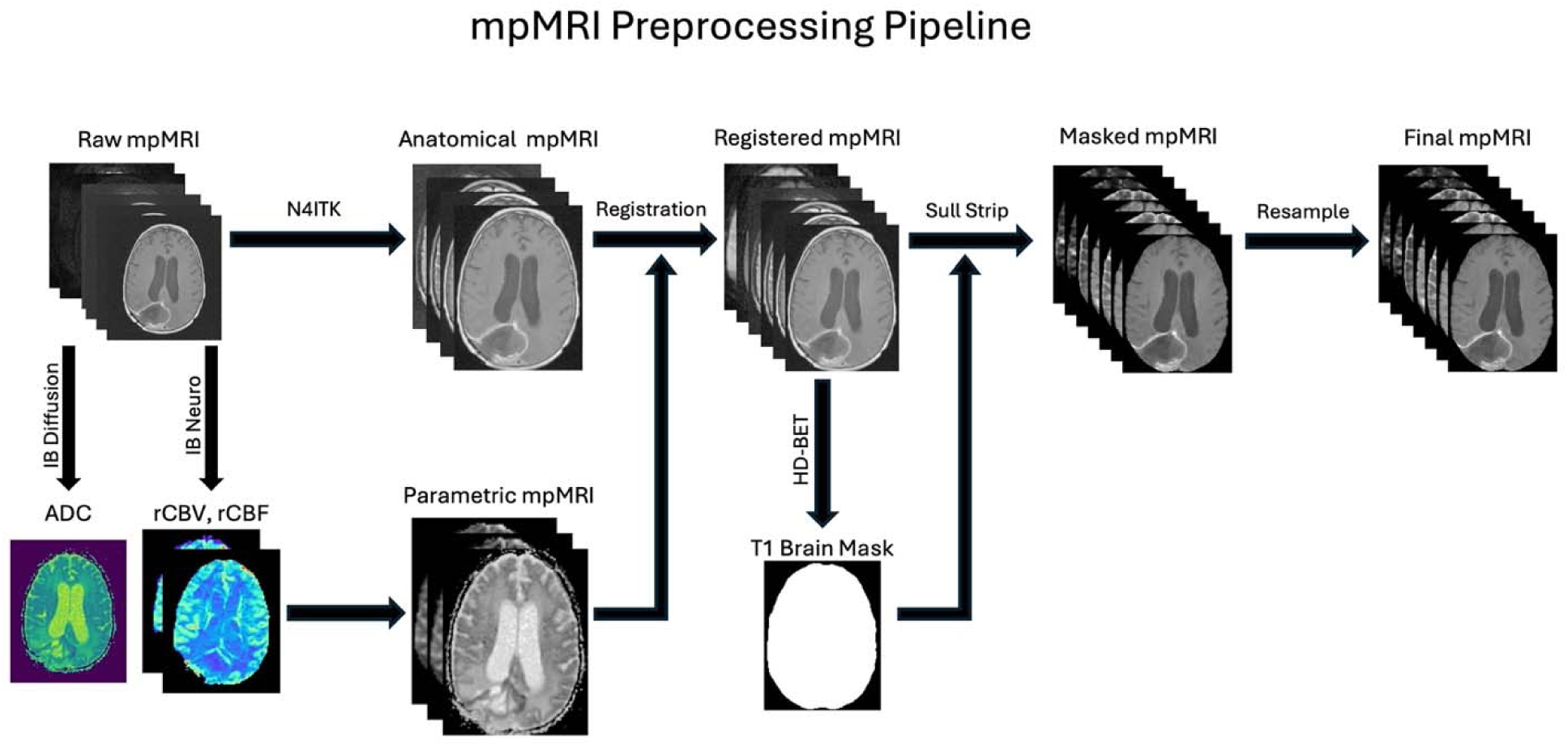
Multiparametric MRI (mpMRI) preprocessing pipeline. Anatomical images (T1-weighted, post-contrast T1-weighted, T2-weighted, and FLAIR) were bias-field corrected using N4ITK and registered to a common anatomical reference. Apparent diffusion coefficient (ADC) maps were generated from diffusion-weighted imaging using IB Diffusion, while relative cerebral blood volume (rCBV) and relative cerebral blood flow (rCBF) maps were generated from dynamic susceptibility contrast (DSC) perfusion imaging using IB Neuro. Parametric maps were registered to the anatomical images. A brain mask was generated from the registered T1-weighted image using HD-BET and applied to remove non-brain tissue. The masked images were then resampled to a common spatial resolution to generate the final mpMRI inputs for model development.

### 2.3 Biopsy Processing

For the glioma patient cohort, biopsy coordinate generation was performed using stereotactic navigation data and intraoperative screen captures of biopsy localization. Biopsy coordinates were derived from the intra-operative screen captures taken of the biopsy site in reference to the Stealth Station navigation scans. Spatial registration between the Stealth Station scans and the mpMRI dataset was performed using modality-matched anatomical images, and biopsy coordinates were mapped into mpMRI space using the resulting transformation. Only biopsies acquired from the non-enhancing regions, as defined by BraTS-derived tumor segmentation maps, were included. Histopathological labels were assigned by an expert neuropathologist and simplified to tumor or non-tumor based on the presence or absence of viable tumor tissue; the non-tumor class corresponds to tissue without histopathological evidence of viable tumor and primarily reflects peritumoral edema.

For the brain metastases cohort, the registered mpMRI dataset and BraTS-derived segmentation of nonenhancing FLAIR-hyperintense regions were used to define candidate sampling regions. This cohort served as a biologically grounded source of non-tumor samples for model development, acting as a proxy for non-infiltrative peritumoral tissue in gliomas. Because brain metastases are typically well-circumscribed and non-infiltrative, with any infiltration generally confined to a minimal distance from the contrast-enhancing lesion, a spatial exclusion strategy was implemented. Specifically, non-enhancing voxels within a 3-mm margin of contrast-enhancing regions were excluded to reduce potential contamination from microscopic infiltration (41). In addition, voxels in non-enhancing regions with fewer than 50% non-enhancing neighbors were removed to avoid sampling from boundary regions. The resulting masked peritumoral region defined the pool of candidate non-tumor sampling locations. From this region, biopsy coordinates were selected using uniform random sampling across patients to achieve class balance.

Three-dimensional ROIs (28x28x28 voxels) centered on biopsy coordinates (**Figure 3**) were extracted from each mpMRI volume serving as model inputs, with corresponding tissue labels serving as model outputs.

**Figure 3.**
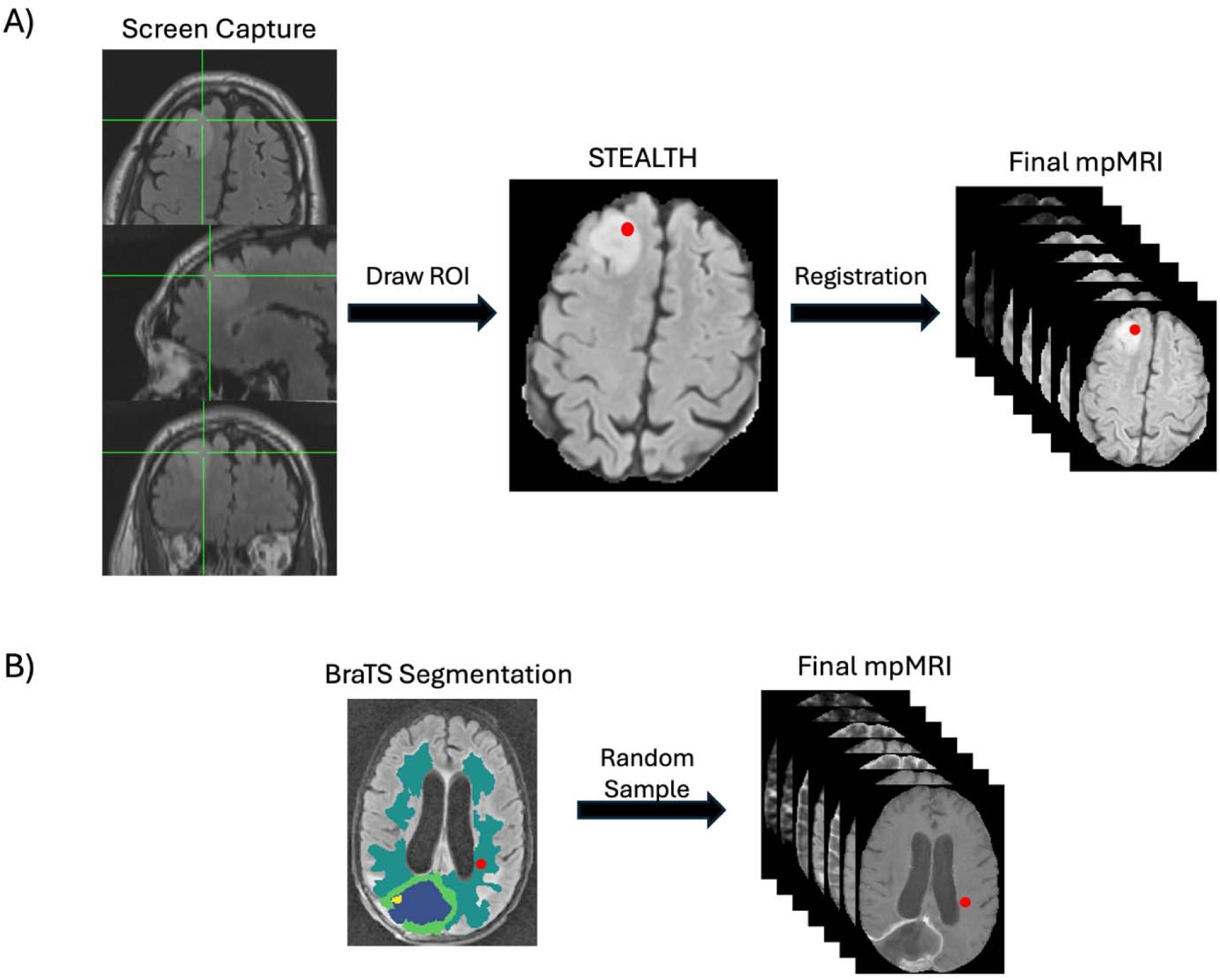
Generation of biopsy-centered regions of interest (ROIs) for model training. (A) For patients with glioma, biopsy coordinates were identified from STEALTH navigation images, with the biopsy target indicated by the green crosshairs. These coordinates defined the center of the ROI, which was mapped onto the corresponding registered multiparametric MRI (mpMRI) datasets using the STEALTH-to-mpMRI spatial transformation. (B) For patients with brain metastases, biopsy coordinates were randomly sampled from the buffered non-enhancing region defined by the BraTS tumor segmentation map. For both cohorts, voxel-centered mpMRI patches (28x28x28 voxels) were extracted as model inputs, with the corresponding tissue labels used as model outputs.

### 2.4 Dataset Composition

Dataset partitioning was performed at the patient level to prevent data leakage between training and evaluation datasets. Glioma and brain metastasis cohorts were partitioned independently to preserve cohort representation across all dataset splits. Twenty percent of patients from each cohort were reserved as an independent test set and excluded from all model development procedures. The remaining patients were used for training and cross-validation (CV), with patient-level separation maintained within each CV fold.

The training mpMRI ROIs underwent on-the-fly augmentation using a custom library of transformations designed for 3D multichannel medical imaging. Augmentation operations included affine transformations, intensity perturbations, histogram-based transformations, additive noise, and posterization, with initial transformation magnitudes selected based on values reported in the literature. Augmentations were applied stochastically at each training epoch to improve model robustness and reduce overfitting and were used exclusively during training.

All mpMRI ROIs were additionally z-score normalized on a per-channel basis using the mean and standard deviation computed from the training dataset.

### 2.5 Infiltrative Tumor Burden Model Development

The infiltrative tumor burden (iTB) model was based on a 3D ResNet (42) backbone with architectural modifications inspired by ConvNeXt (43), incorporating design principles from vision transformers within a convolutional framework. The model processes 3D multichannel voxel-centered patches (28 x 28 x 28) extracted from mpMRI volumes, enabling voxel-wise classification. The overall architecture is illustrated in **Figure 4**.

**Figure 4.**
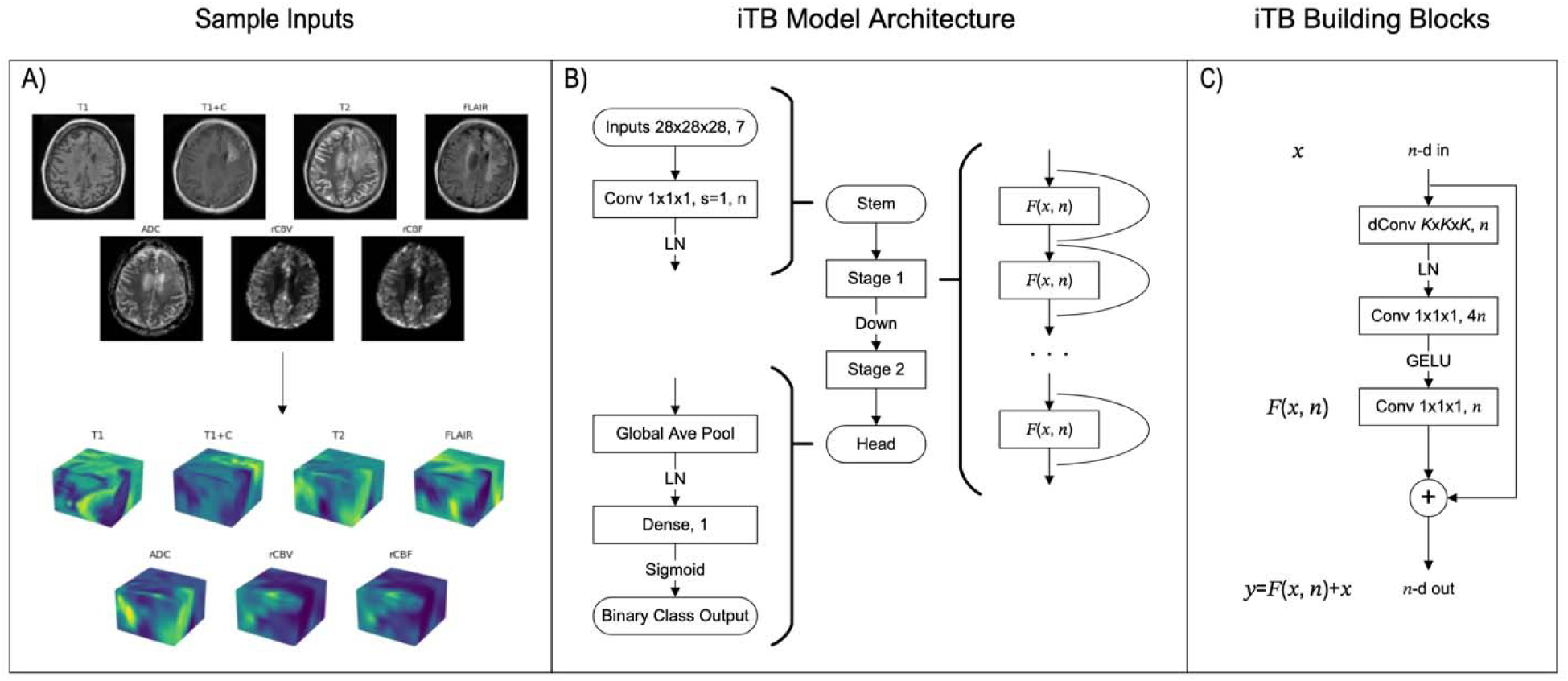
Overview of the infiltrative tumor burden (iTB) model. The model takes as input 3D voxel-centered, multi-channel patches (28x28x28 voxels) extracted from mpMRI volumes (A). These patches are processed by the iTB model whose architecture is based on a ConvNeXt-modified 3D ResNet neural network (B). The constituent network building blocks are illustrated in (C).

The network consists of a stem, two hierarchical feature extraction stages with residual building blocks, and a classification head. Compared to standard ResNet architectures, the number of stages was reduced and the down sampling strategy was adapted to accommodate the smaller input patch size. Each stage comprises multiple residual blocks incorporating depth wise 3D convolutions, inverted bottleneck design, and layer normalization. GELU activations are applied within each block following the ConvNeXt design paradigm. Spatial down sampling between stages is performed using dedicated stride convolution layers. The model head consists of global average pooling followed by layer normalization and a fully connected layer with sigmoid activation for binary voxel-wise classification.

Binary cross-entropy was used as the loss function. Regularization was achieved using weight decay, label smoothing, and stochastic depth. Model training and evaluation were performed using group-wise 3-fold CV to prevent patient-level data leakage. Brain metastasis samples used for non-tumor reference were re-sampled every 10 epochs during training to reduce overfitting to a fixed negative set and improve variability in negative class representation.

Hyperparameter optimization was conducted within this framework using random search, with the optimal configuration selected based on the area under the receiver operating characteristic curve (AUC) computed from pooled out-of-fold predictions across held-out folds.

A secondary optimization procedure was performed to refine augmentation magnitudes. The best-performing architectural hyperparameters identified during the previous optimization procedure were fixed throughout this process. To limit computational cost, augmentation magnitudes were optimized using a greedy local-search strategy rather than exhaustive exploration of the augmentation parameter space. Baseline performance was first established using augmentation magnitudes commonly reported in the literature. At each iteration, an augmentation type was randomly selected from the active candidate set and its magnitude was perturbed by a fixed increment. The model was retrained and evaluated within the CV framework using pooled out-of-fold AUC as the optimization metric. If performance improved, additional perturbations in the same direction were evaluated iteratively until no further improvement was observed, with the best-performing magnitude retained. If the initial perturbation failed to improve performance, an equivalent perturbation in the opposite direction was evaluated. Augmentation types for which neither positive nor negative perturbations improved performance were removed from the candidate set. The process continued until no augmentation types remained eligible for further optimization.

The model was implemented in TensorFlow (version 2.16.1; Google, Mountain View, CA, USA) and trained on NVIDIA Ampere A40 and Tesla V100 GPUs.

### 2.6 Analysis

CV performance was computed from pooled out-of-fold predictions aggregated across held-out folds, ensuring patient-level independence between training and validation data. The final model configuration, including architectural hyperparameters and augmentation settings selected during optimization, was subsequently evaluated on an independent test set comprising 20% of patients, which was not used during model development or model selection.

Model performance was primarily assessed using AUC. Secondary metrics assessed included accuracy, sensitivity, specificity, positive predictive value (PPV), and negative predictive value (NPV). Uncertainty in AUC estimates was quantified using non-parametric patient-level bootstrap resampling (1,000 iterations) to derive 95% confidence intervals. Statistical comparisons between baseline and augmentation-optimized models were performed on the independent test set using a one-sided DeLong test for paired ROC curves (p < 0.05) to assess whether augmentation optimization improved discriminative performance. The test was applied to predictions generated from the same patients, accounting for correlation between model outputs.

For voxel-wise interpretability and initial feasibility assessment, iTB probability maps were generated for representative cases from the independent test set, including two brain metastasis cases (serving as negative controls) and one glioblastoma case (serving as a positive control where infiltration is expected). Inference was performed in T1-aligned space within regions constrained by the BraTS-derived non-enhancing mask, yielding voxel-wise probability estimates of tumor infiltration.

To improve robustness, ensemble inference was performed across the three independently trained iTB models. Voxel-wise probabilities were averaged across models to produce a single continuous probability map, reducing model variance while preserving spatial uncertainty. Final binary segmentation maps, distinguish tumor from non-tumor, were obtained by applying a global threshold determined using the Youden J statistic derived exclusively from CV ROC analysis.

All statistical analyses were performed in Python (version 3.10).

## 3.0 RESULTS

### 3.1 Dataset

Following application of inclusion criteria, 80 glioma patients and 24 brain metastasis patients were available for analysis. Among glioma patients, 266 biopsy-localized samples with corresponding intraoperative screen-capture localization were identified. Of these, 121 biopsies were acquired from non-enhancing regions of 56 patients and were eligible for machine learning model development. The remaining 176 biopsies originated from contrast-enhancing regions or regions that could not be classified as either enhancing or non-enhancing and were therefore excluded from model development. The median interval between imaging and surgery was 11 days.

The brain metastasis cohort consisted of 26 patients with a range of primary tumor histologies. For this cohort, non-tumor training samples were generated by randomly sampling up to four non-overlapping ROIs from buffered non-enhancing, peritumoral regions as defined in the Methods. Sampling locations were dynamically resampled during training to increase spatial diversity, whereas fixed coordinates were retained for validation and testing. Details of the dataset are provided in **Table 1**.

**Table 1.** Summary of the cohorts used for iTB model development and evaluation. Demographic characteristics, imaging-to-surgery intervals, and biopsy sample counts and histopathological classifications are summarized for the glioma and brain metastasis cohorts. Glioma cases are additionally stratified by diagnosis. Continuous variables are reported as mean ± standard deviation where indicated. Tumor-positive and tumor-negative samples are classified according to histopathological assessment. NA, not applicable.

| Diagnosis | No. Patients | Sex (Male : Female) | Age at Imaging (years; ave ± std) | Time between Imaging and Surgery (days; ave ± std) | No. Samples | Sample Pathology (tumor +:tumor -) |
| --- | --- | --- | --- | --- | --- | --- |
| Glioma Cohort |  |  |  |  |  |  |
| Astrocytoma | 22 | 17:5 | 41 ± 13 | 11 ± 7 | 67 | 60:7 |
| Glioblastoma | 30 | 15:15 | 55 ± 11 | 7 ± 5 | 46 | 38:8 |
| Oligodendroglioma | 4 | 1:3 | 32 ± 5 | 5 ± 4 | 8 | 7:1 |
| Cohort Total | 56 | 33:23 | 48 ± 14 | 9 ± 6 | 121 | 105:16 |
| Brain Metastasis Cohort | 26 | 5:21 | 61 ± 11 | NA | NA | NA |
| Dataset Total | 82 | 38:44 | 52 ± 12 | NA | NA | NA |

All training samples were constructed as 3D multichannel voxel-centered patches containing co-registered T1, T1+C, T2, FLAIR, rCBV, rCBF, and ADC image channels as described in the Methods section. All reported performance metrics were evaluated exclusively on an independent test set, defined as a 20% patient-level hold-out subset of glioma and brain metastasis cases that was excluded from model training, hyperparameter tuning, and model selection.

### 3.2 Model Performance

Hyperparameter optimization was constrained to 200 randomly sampled configurations, with training times ranging from approximately 1 to 4 hours per configuration. The model configuration yielding the highest pooled CV AUC is presented in **Table 2**.

**Table 2.** Optimized hyperparameters of the three-dimensional deep learning model developed to estimate infiltrative tumor burden (iTB) within non-enhancing, FLAIR-hyperintense lesions. . The table summarizes the optimal values selected through hyperparameter optimization, including training configuration, regularization, learning rate scheduling, and network architecture parameters.

| Hyperparameter | Optimal Value |
| --- | --- |
| Label Smoothing | 0.2 |
| Optimizer | RMSProp |
| Weight Decay | 0.01 |
| Momentum | 0.9 |
| LR Decay Function | cosine |
| Warmup Epochs | 0.10% |
| Training Epochs | 50 |
| Layer Scaling Initialization | 0.1 |
| SD Drop Rate | 0.3 |
| Stage Ratio | [3,9] |
| Network Width | 32 |
| DW Convolutional Filter Size | 3 |

The default and optimized augmentation configurations are presented in **Table 3**. Following optimization, most augmentation magnitudes were reduced relative to baseline settings following optimization. Three-dimensional rotation magnitude increased slightly, whereas blur and equalization were reduced to zero following optimization.

**Table 3.** Default and optimized data augmentation configurations used during training of the iTB model. The table summarizes the augmentation methods and their corresponding magnitude settings before and after optimization. Geometric transformations included 3D flipping, rotation, shear, and translation, while intensity and image-quality transformations included Gaussian blur, sharpening, channel blending, autocontrast, equalization, additive noise, and posterization. Optimal settings were selected through augmentation hyperparameter optimization to improve model generalization while preserving relevant imaging characteristics.

| Table 3: Default and Optimized Augmentation Configurations |  |  |
| --- | --- | --- |
| Augmentation Method | Default Magnitude | Optimal Magnitude |
| 3D Flip<br>(random subset of xy, xz, and yz planes) | 50% probability | 50% probability |
| 3D Rotation<br>(random axis; random angle between 0 and indicated value) | Angle = 22.5° | Angle = 27° |
| 3D Shear<br>(random axis and random direction; random magnitude between 0 and indicated value) | Shear factor = .15 | Shear slope = .15 |
| 3D Translation<br>(random direction; random magnitude between 0 and indicated value) | 15% edge length | 15% edge length |
| Blur<br>(blending with volume that underwent convolution with Gaussian blur kernel; random percentage between 0 and indicated value) | 50% | 20% |
| Sharpness<br>(blending with volume that underwent convolution with 27-point stencil kernel; random percentage between 0 and indicated value) | 50% | 30% |
| Color<br>(blending with volume that is an equally weighted sum of all channels; random percentage between 0 and indicated value) | 50% | 0% |
| AutoContrast<br>(random cufoff percentage between 0 and indicated value) | 5% cutoff | 4% cutoff |
| Equalize | 20% probability | 0% probability |
| Additive Noise<br>(random magnitude between 0 and indicated value) | 50% | 90% |
| Posterize<br>(random bit-shift between 0 and indicated value) | Bit-shift = 4 | Bit-shift = 4 |

Model performance in both pooled CV and independent test-set evaluations is reported in **Table 4**. On the test set, accuracy increased from 74.5% to 85.5%, and AUC increased from 0.859 to 0.942 following augmentation optimization. Improvements were also observed in pooled CV performance, with accuracy increasing from 83.0% to 87.5% and AUC increasing from 0.884 to 0.928.

**Table 4.**
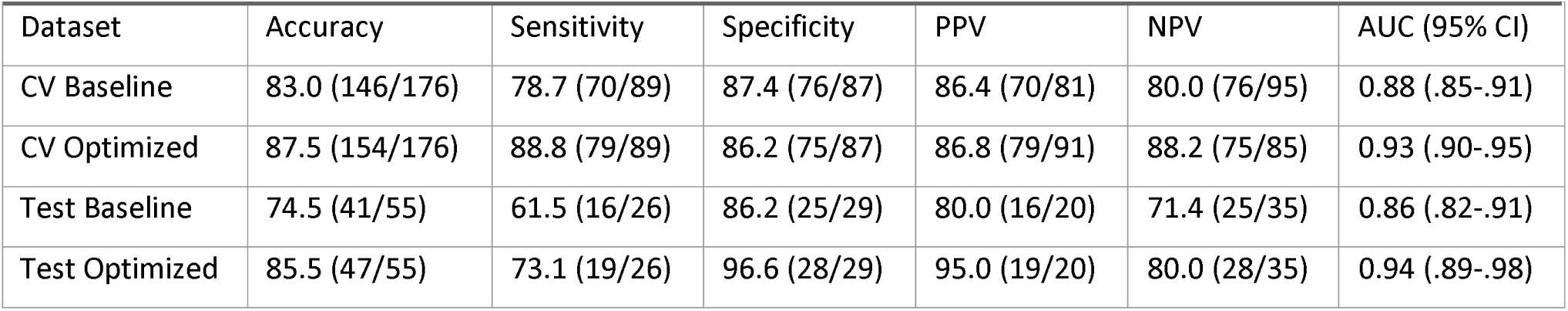
Performance of the baseline and optimized infiltrative tumor burden (iTB) models in cross-validation (CV) and held-out test datasets. Classification performance is reported as accuracy, sensitivity, specificity, positive predictive value (PPV), and negative predictive value (NPV), with corresponding numerators and denominators in parentheses. The area under the receiver operating characteristic curve (AUC) is reported with 95% confidence intervals (CIs). Optimization improved overall accuracy and AUC in both datasets, with the largest gains in test-set sensitivity and specificity.

ROC curves for baseline and optimized models are shown in **Figure 5**. Improvement in test-set AUC was statistically significant based on a one-sided DeLong test (p = 0.008).

**Figure 5.**
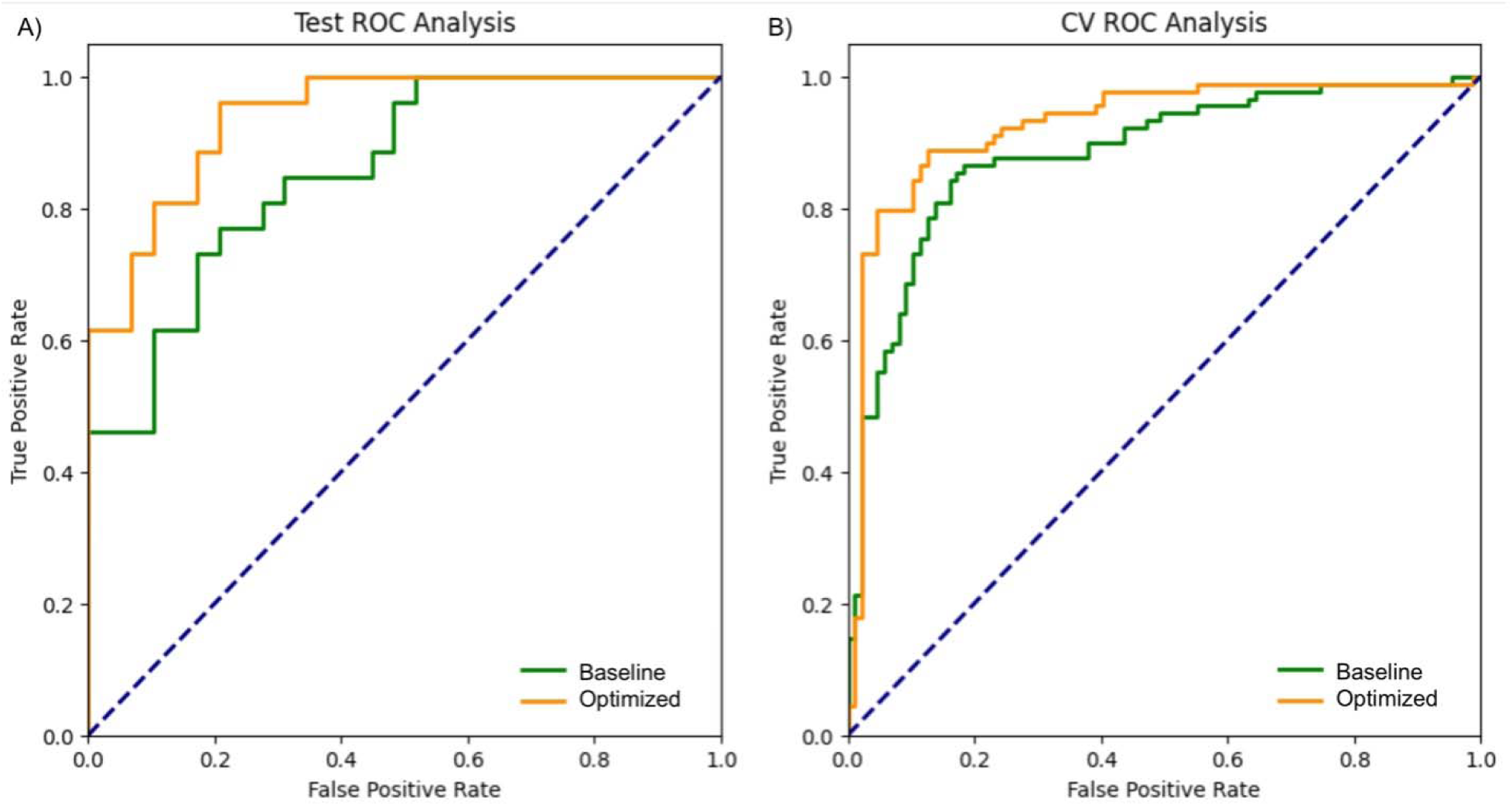
Receiver operating characteristic (ROC) curves comparing the baseline and optimized models using an independent test set (A) and pooled cross-validation (B). In both evaluations, the optimized data augmentation strategy improved model performance. For the independent test set, accuracy increased from 74.5% to 85.5%, while the area under the ROC curve (AUC) increased significantly from 0.859 to 0.942 (p=0.008) demonstrating improved generalizability of the optimized model. For the pooled CV, accuracy increased from 83.0% to 87.5%, and the AUC increased from 0.884 to 0.928.

### 3.3 Ensemble Inferencing and iTB Map Generation

Ensemble inference was performed to generate voxel-wise iTB probability maps in the independent test set, including two brain metastasis cases and one glioblastoma case. Predictions were generated in T1-aligned space within the non-enhancing mask, yielding voxel-wise probability estimates of tumor infiltration.

For each voxel, probabilities from the three independently trained models were averaged to produce a single ensemble probability map. This probability-level fusion preserved continuous spatial structure prior to thresholding.

Binary maps were derived by applying a fixed decision threshold to the ensemble probability maps. This operating threshold was selected from CV using the Youden J statistic (J = 0.572) and applied uniformly across all test cases to separate voxels classified as infiltrative tumor from those consistent with non-tumor tissue.

Representative iTB maps are shown in **Figure 6** for two brain metastasis cases and one glioblastoma case from the independent test set. Voxel-wise predictions range from 0.0 to 1.0, where values near 0 indicate low probability of tumor infiltration and values near 1 indicate high probability.

**Figure 6.**
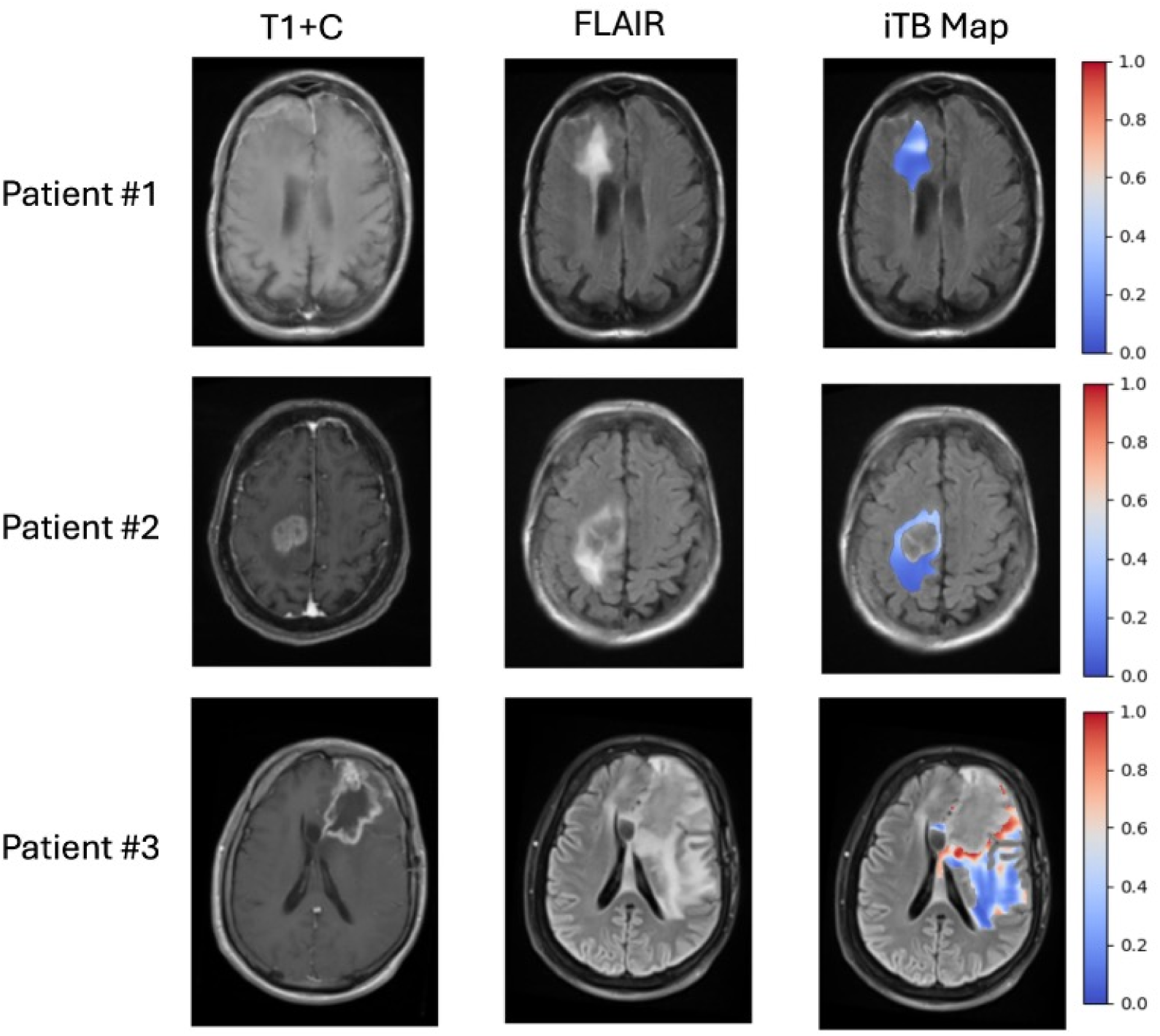
Representative infiltrative tumor burden (iTB) maps from two patients with brain metastases and one patient with glioblastoma. Voxel-wise iTB predictions range from 0.0 to 1.0, with values near 0 indicating a low probability of tumor infiltration and values near 1 indicating a high probability. Consistent with the expected growth patterns of brain metastases, non-enhancing regions showed predominantly low iTB values. In contrast, the glioblastoma case demonstrated elevated iTB values extending beyond the enhancing tumor margin into surrounding non-enhancing tissue, consistent with more diffuse infiltrative nature of glioblastoma.

In both brain metastasis cases (Patients #1 and #2), predicted tumor probability within the non-enhancing region remained largely below the classification threshold, with only small, localized regions approaching threshold values. These regions were spatially isolated and did not form contiguous clusters exceeding the classification threshold.

In contrast, the glioblastoma case (Patient #3), for which infiltrative tumor growth is expected, demonstrated broader and more spatially contiguous regions of elevated probability within the non-enhancing mask, consistent with infiltrative signal on iTB mapping.

The average runtime for iTB map generation was approximately 10 minutes per case when ensemble models were executed in parallel followed by fusion, with increased processing time observed for larger non-enhancing regions.

## 4.0 DISCUSSION

A new biomarker of infiltrative tumor burden within non-enhancing, hyperintense FLAIR lesions was developed using mpMRI and deep learning with spatially matched diagnostic tissue samples from brain tumor patients. Additionally, a greedy local-search optimization procedure was introduced to fine-tune the magnitudes governing data augmentation during training.

Compared to previous studies with similar objectives, the iTB model demonstrates improved performance in terms of accuracy and AUC while addressing common limitations in study design, including limited dataset size, reliance on indirect or incomplete labeling strategies, and lack of independent test evaluation. These improvements are enabled by the use of a relatively large histologically validated dataset with spatially matched stereotactic biopsy samples and a fully independent test cohort, providing a more robust estimate of model performance and reducing reliance on cross-validation-based reporting, which may overestimate generalization. In addition, performance gains were further supported by an augmentation optimization strategy, which systematically refined transformation magnitudes rather than relying on fixed literature-derived settings, contributing to improved generalization and highlighting the importance of dataset-specific tuning of augmentation strategies in medical imaging applications.

Initial iTB maps demonstrate qualitative agreement with expected biological behavior of different tumor types, supporting the feasibility of voxel-wise infiltration mapping. In glioblastoma, where infiltrative growth is well established, regions of elevated iTB values extend throughout the non-enhancing region, consistent with diffuse tumor spread. In contrast, brain metastasis cases show predominantly low iTB probabilities within the non-enhancing regions, reflecting their largely non-infiltrative growth pattern. Together, these findings provide preliminary support for the model’s ability to distinguish between infiltrative and non-infiltrative phenotypes.

Although the present study includes a relatively large dataset for spatially matched neuro-oncologic imaging studies, continued expansion remains necessary to further improve model robustness and generalizability. A key limitation relates to the inherently non-uniform nature of stereotactic biopsy sampling, which is clinically directed rather than systematically distributed across non-enhancing regions. This contributes to an imbalance in available tissue labels, particularly for non-tumor samples within glioma cases, and motivated the inclusion of a brain metastasis cohort to better represent peritumoral tissue environments where vasogenic edema is present without tumor cells. A limitation of stereotactic biopsy–based labeling is the mismatch in spatial resolution between biopsy localization and imaging, which can introduce minor uncertainty in assigning labels to individual voxels.

While the dataset was acquired within a single health system comprising multiple sites, external generalizability to other institutions and geographic regions remains to be established.

Due to dataset size constraints, augmentation optimization was performed within the same cross-validation framework used for architectural hyperparameter selection, rather than a fully nested cross-validation design. As a result, performance estimates may retain a modest degree of optimism related to repeated model selection within the validation framework.

Another limitation is the retrospective, single-health system nature of the dataset, which may restrict generalizability to external imaging protocols and patient populations. Although this dataset exceeds many prior studies in this domain, it remains modest in scale compared to large-scale datasets in non-medical machine learning applications. Furthermore, the current model relies on a multi-sequence mpMRI protocol that includes seven imaging-derived inputs. While this improves predictive performance, it may limit applicability in settings where full multiparametric acquisition is not routinely available. A sensitivity analysis is therefore planned to evaluate reduced input configurations and identify minimal imaging requirements for robust inference.

From a clinical perspective, voxel-wise mapping of infiltrative tumor burden within non-enhancing peritumoral regions provides a framework for refining how infiltrative disease is represented in treatment planning. In surgical contexts, this may support more informed decisions regarding the extent of resection beyond contrast-enhancing tumor by identifying spatial gradients of likely infiltration. In radiotherapy planning, iTB maps may contribute to more precise definition of target volumes by distinguishing regions of higher versus lower likelihood of tumor infiltration within FLAIR abnormality, potentially improving targeting specificity while maintaining oncologic coverage. More broadly, this framework introduces a quantitative representation of infiltration that may support more consistent interpretation of peritumoral disease burden across time and between patients, particularly in the context of treatment response assessment.

## Data Availability

Data generated or analyzed during the study are available from the corresponding author by request.

## Acknowledgements

We would like to acknowledge [Redacted] and [Redacted] for their substantial contribution to this work, specifically recognizing their efforts in tissue procurement and record-keeping.

During the preparation of this work, the authors used CoPilot for assistance with editing, mostly in the form of rephrasing text to make it clearer and more concise. The authors reviewed and edited the output as needed and take full responsibility for the content of the published article.

